# Osteoporosis and its associated factors among patients attending Manakamana hospital, Chitwan

**DOI:** 10.1101/2023.07.21.23293006

**Authors:** Shankar Dhakal, Kalpana Sharma, Kishor Adhikari, Alisha Joshi, Sunita Poudyal

**Author notes:** **Correspondence** Dr. Kishor Adhikari, Professor, School of Public Health and Dept. of Community Medicne, Chitwan Medical College, Nepal.

## Abstract

**Background:** Osteoporosis is most common skeletal disorders that weaken bones and increase their susceptibility to fractures. It is becoming an urgent and serious global epidemic.Early diagnosis and treatment are essential to reduce morbidity and mortality associated with it. This study aimed to find out the prevalence of osteoporosis among patients attending at Manakamana Hospital, Bharatpur, Chitwan.

**Methods:** A descriptive, cross-sectional study was adopted and 623 patients attending at orthopaedic outpatients department (OPD) of Manakamana Hospital were selected using consecutive sampling technique. Data were collected from 15^th^ October 2021 to 15^th^ April, 2022, by using interview schedule, chart review and BMD measurement through calcaneal ultrasonography. Ethical approval was obtained from NHRC-IRC prior to study procedures. Obtained data were analysed using descriptive statistics and association between the variables were measured using chi-square test.

**Results:** The mean age of the patients was 43.5 (±14.26) years. Nearly half (44%, n=274) were middle aged adults, 59.7% were female and 56.0% were involved in agriculture and household chores. Nearly half of the patients (45.7%) were overweight/ obese, 7.9% were smokers and 13.5% had habit of alcohol use. Osteopenia or low bone density was detected in 58.9% patients and 19.4% had osteoporosis. The prevalence of osteoporosis was significantly associated with age group (p=<0.001) and educational status (p=0.013) of the patients.

**Conclusions:** Osteoporosis and osteopenia are prevalent in patients attending in the hospital. Hence, awareness, early screening, treatment is necessary for the hospital attended patients to minimize the risk of fracture and the consequences associated with it.

## Introduction

Osteoporosis is most common skeletal disorders that weaken bones and increase their susceptibility to fractures. It is becoming an urgent and serious global epidemic which accounts 18.3% of the world. Asia has higher prevalence of osteoporosis (24.3%) than the USA, and Australia but lower prevalence than Africa and Europe [1]. Currently it is estimated that approximately 500 million people worldwide suffer from this disease [2,3]. Osteoporotic fractures are characterized as fractures associated with low bone mineral density (BMD) [4]. Quantitative Ultrasonography (QUS) is a quick and reliable method for identifying osteoporosis in both men and women in female in developing nations [5].

In Nepalese population, the prevalence of osteoporosis is high [6,7,8]. Unfortunately, it is often undiagnosed until a fracture occurs. Osteoporotic fractures have significant, long-lasting impacts on mortality, increase in functional limitations and decreases in quality of life, and place a significant financial and human resource burden on healthcare systems [9] Few studies have been conducted in Nepalese population [6,7,8,10]. Hence, this study aimed to assess the prevalence of osteoporosis among patients attending Manakamana Hospital, Chitwan.

## METHODS

A descriptive cross-sectional study was conducted among the patients attending Orthopaedic Out Patients Departments of Manakamana Hospital, Chitwan to collect the information regarding prevalence of osteoporosis. Data were collected from 15^th^October 2021 to 15^th^April, 2022. Consecutive sampling technique was used to select the study sample. There were total 623 patients who met the study criteria and attended in the OPD during the study periods so all of them were taken as study sample. This study included patients aged 20 years and above, visiting the hospital during data collection period and willing to participate in the study whereas those patients who had previous history of fracture, pregnant and severely ill or unable to communicate were excluded from the study.

After getting ethical approval from Nepal Health Research Council Institutional Review Committee (NHRC-ERB, Ref No.3137) and data collection permission from the selected hospital, data were collected by the researchers using structured interviews schedule, bio-physiological measurementand records reviews.First, patients were identified from their records file. The purpose of the study was explained and written informed consent was taken. Data related to socio-demographic, personal habit and other related information were collected using a structured interview schedule. Then weight and height were measured through weighing scale and stadimetry. After that, bone mass density (BMD) was measured at calcaneus (heel bone) using portable Ultrasound bone Densimeter (SONOST 2000). The measurement was done on the right heel for all patients. The examinations were performed by a single trained professional. The apparatus was calibrated daily.

The collected data were coded and entered into IBM SPSS^®^ software version 20 for window. Continuous variables were presented as mean, standard deviation, median, and interquartile range while categorical variables were presented as the frequency and percentage. The prevalence of osteoporosis was estimated through WHO criteria for osteoporosis (normal BMD if T-score >-1 SD, osteopenia if T-score is -1 SD to -2.5 SD, and osteoporosis if T-score <-2.5 SD). BMI was calculated as weight in kilograms/ height in meter square. Chi-square test was applied to measure the association between prevalence of osteoporosis and selected variables. All the statistical significance was set at p<0.05.

## RESULTS

Out of 623 respondents, 274(44.0%) patients were middle aged adults, 375 (59.7%) were female, 557 (89.4%) had completed secondary and above level of education, and 349 (56.0%) were involved in agriculture and household chores. Regarding personal habit, 49 (7.9%) were smokers and 84 (13.5%) had habit of alcohol use. Out of total respondents, 285 (45.8%) were overweight and obese and few had high systolic BP 40 (6.4%) and diastolic BP 70 (11.2%)(Table1).

**Table 1:** Socio-demographic and Personal Habit of Respondents.

| **n=623** |  |  |
| --- | --- | --- |
| **Characteristics** | **Number** | **Percentage** |
| **Age (completed in years)** |  |  |
| Young (20-39 year) | 262 | 42.1 |
| Middle (40-59 year) | 274 | 44.0 |
| Elderly ($\geq 60$ year) | 87 | 14.0 |
| *Mean (SD)=43.5($\pm 14.26$), Min-20 year ,Max-99 year* |  |  |
| **Sex** |  |  |
| Male | 251 | 40.3 |
| Female | 375 | 59.7 |
| **Education** |  |  |
| Illiterate | 37 | 5.9 |
| Basic | 29 | 4.7 |
| Secondary | 273 | 43.8 |
| Bachelor and above | 284 | 45.6 |
| **Occupation** |  |  |
| Agriculture and Household work | 349 | 56.0 |
| Service | 170 | 27.3 |
| Business | 78 | 12.5 |
| Others (Retired, Students) | 26 | 4.2 |
| **Smoking Habit** |  |  |
| Yes | 49 | 7.9 |
| No | 574 | 92.1 |
| **Alcohol Habit** |  |  |
| Yes | 84 | 13.5 |
| No | 539 | 86.5 |
| **BMI Status** |  |  |
| Underweight (<18.5) | 42 | 6.7 |
| Normal (18.5-24.9) | 296 | 47.5 |
| Overweight (25.0-29.9) | 185 | 29.7 |
| Obesity (30 and above) | 100 | 16.1 |
| **Systolic BP** |  |  |
| Normal (<120 mm of Hg) | 277 | 44.5 |
| Pre-hypertensive (120-139 mm of Hg) | 306 | 49.1 |
| Hypertensive (>140 mm of Hg) | 40 | 6.4 |
| **Diastolic BP** |  |  |
| Normal (<80 mm of Hg) | 307 | 49.3 |
| Pre-hypertensive (80-89 mm of Hg) | 246 | 39.5 |
| Hypertensive (>90 mm of Hg) | 70 | 11.2 |

Out of 623 respondents, 367 (59.9%) respondents had osteopenia or low bone density and 121 (19.4%) respondents had osteoporosis (Table 2).

**Table 2:** Prevalence of Osteoporosis among the Respondents.

| **Osteoporosis Status** | **Frequency** | **Percentage** |
| --- | --- | --- |
| Normal bone density (T-score is equal to or above -1 S.D). | 135 | 21.7 |
| Osteopenia or low bone density (T-score ranges between -1.1 S.D. and -2.5 S.D), | 367 | 58.9 |
| Osteoporosis (T-score is equal to or below -2.5 S.D). | 121 | 19.4 |
| Total | 623 | 100 |

Prevalence of osteoporosis was significantly associated with age (*p*=<0.001) and education (*p*=<0.013) of the respondents whereas other variables were not associated with it (Table 3).

**Table 3:** Association between Prevalence of Osteoporosis and Selected Variables.

**n=623**
| **Variables** | **Prevalence Osteoporosis** |  |  | **x<sup>2</sup>** | **p- value** |
| --- | --- | --- | --- | --- | --- |
|  | **Normal** | **Osteopenia** | **Osteoporosis** |  |  |
| **Age Group** |  |  |  |  |  |
| Young | 62 (23.7) | 170 (64.9) | 30 (11.5) | 70.963 | <0.001 |
| Middle | 65 (23.7) | 163 (59.5) | 46 (16.8) |  |  |
| Elderly | 8 (9.2) | 34 (39.1) | 45 (51.7) |  |  |
| **Sex** |  |  |  |  |  |
| Male | 63 (25.1) | 140 (55.8) | 48 (19.4) | 3.002 | 0.223 |
| Female | 72 (19.4) | 227 (61.0) | 73 (19.6) |  |  |
| **Education** |  |  |  |  |  |
| Illiterate | 9 (24.3) | 14 (37.8) | 14 (37.8) | 16.140 | **0.013** |
| Basic | 5 (17.2) | 20 (69.0) | 4 (13.8) |  |  |
| Secondary | 71 (26.0) | 157 (57.5) | 45 (16.5) |  |  |
| Bachelor and above | 50 (17.6) | 176 (62.0) | 58 (20.4) |  |  |
| **Occupation** |  |  |  |  |  |
| Business | 21 (26.9) | 46 (59.0) | 11 (14.1) | 4.102 | 0.663 |
| Farmer/agriculture | 70 (20.1) | 204 (58.5) | 75 (21.5) |  |  |
| Service | 39 (22.9) | 100 (58.8) | 31 (18.2) |  |  |
| Other<br>(retired/student) | 5 (19.2) | 17 (65.4) | 4 (15.4) |  |  |
| **Smoking Habit** |  |  |  |  |  |
| No | 119 (20.7) | 346 (60.3) | 109 (19.0) | 5.798 | 0.050 |
| Yes | 16 (32.7) | 21 (42.9) | 12 (24.5) |  |  |
| **Alcohol Habit** |  |  |  |  |  |
| No | 111 (20.6) | 323 (59.9) | 105 (19.5) | 2.702 | 0.259 |
| Yes | 24 (28.6) | 44 (52.4) | 16 (19.0) |  |  |
| **BMI Status** |  |  |  |  |  |
| Underweight | 11 (26.2) | 17 (40.5) | 14 (33.3) | 7.163 | 0.128 |
| Normal | 63 (21.3) | 180 (60.8) | 53 (17.9) |  |  |
| Overweight/obese | 61 (21.4) | 170 (59.6) | 54 (18.9) |  |  |

## DISCUSSION

This study was aimed to assess the prevalence of osteoporosis for the effective prevention and management of osteoporosis and osteopenia in patients. In this study, prevalence of osteoporosis and osteopenia were 19.4% and 59.9% respectively according to Bone Mass Density (BMD) scoring through QUS. It is consistent with a study from central part of Nepal [6], global data of osteoporosis [1,11] and Hongkong [12]. Whereas it was lower than the studies from Kathmandu, Nepal [10], Panjab, India [13] and United States [14]. The discrepancy rates of osteoporosis in the studies might be due to different nature of population included in the study, sample size, and life style of study participants. Further, our finding was also higher than the study done in Nepal [7] which might be due to use of different measuring tool for the osteoporosis.

Osteoporosis varies among population to population according to their age [15]. In our study osteoporosis is higher among middle aged and elderly population compared to young adult. It is similar with other studies conducted in Nepal [6,7,10,14,16} and China [17]. This might be due to the facts that aging is associated with decreases in the growth hormone secretion from the anterior pituitary and decreased systemic and local skeletal production of IGF-1 and IGF-2, growth factor binding proteins which may contribute to age-related bone loss [18].

The present study found that the prevalence of osteoporosis is higher among participants with lower education levels compared to people with higher education level and this is statistically significant. This finding agreed with the studies conducted in Northwestern of China [16] and morocco [19] which reported the significant association between educational level and risk of osteoporosis. This finding is in line with the western countries finding in which low educated women are more prone to low density bone and osteoporosis than high educated women [20,21,22].

Although the public mislabelled osteoporosis with women, it can affect both men and women equally^23^ and our finding is consistent with it showing that there was no significance difference on the prevalence of osteoporosis between women and men. However, other studies from Italy [24] and China [17,25] have shown the higher incidence of osteoporosis in women compared to men. Likewise, a systematic review and meta-analysis [1] revealed higher prevalence of osteoporosis among women in the world compared to men (women-23.2%, men-11.7%). Further, evidence showed that the bone thinning begins in both men and women between the ages of 35 and 40. In contrast to women, who experience extra bone loss associated with oestrogen insufficiency during peri-menopausal and post-menopausal periods, men are seen to experience a minor longitudinal bone loss throughout life [26].

In our study, we found that there was no significant association between the prevalence of osteoporosis with the occupation, smoking status and alcohol consumption of the patients. In contrast to this, findings from Nepal [10], USA [14] and China [16] have shown that smoking [10,14,16], alcohol consumption, physical activity {16] and daily dietary calcium intake [10] as other modifiable risk factors of osteoporosis. The difference might be due to nature of population, and sites of BMD measurement. Further, our study did not measure the detail of these variables.

Several studies had supported that the higher body mass index (BMI) is a protective factor for osteoporosis [10,13,19] BMI and BMD are positively associated [8,27]. Patients with high BMI had a higher BMD and a lower risk of osteoporosis than those with a normal BMI [28]. Our study revealed that BMI is not statistically associated with the prevalence of osteoporosis. Unlike this finding, other studies [6,14,16] revealed the significant negative relationship between BMI and osteoporosis. The difference in findings might be due to difference populations and the settings.

This study has some limitations and strength to note. Owing to cross-sectional study design, this was unable to establish cause and effects relationship. Additionally, patients were selected from orthopaedic OPD of a hospital; general prevalence could be different. Furthermore, inability to explore the data on other risk factors like co-morbidities, drugs, and nutritional status could be another limitation of this study. However, our findings are reflective of prevalence of osteoporosis among patients attending to orthopaedic OPD because of relatively large sample size. Also using portable DEXA scan aligns our research with current international osteoporosis foundations guidelines for evaluating osteoporosis. These estimates could be helpful in determining priorities, formulating policies, and allocating resources for osteoporosis prevention and treatment. Large scale, multi-centric, randomized sampling study is needed to minimize the bias and establish the real burden of the problem.

## CONCLUSIONS

Patients’ population visiting hospital is at high risk of osteoporosis and osteopenia. Higher age and lower education status patients are at increased risk of osteoporosis. There is need of early detection of osteoporosis among hospital attended patients to minimize the possible fracture risk and consequences. Further, awareness programs are needed to these risk groups to minimize the possible fracture risk and consequences.

## Data Availability

Data are fully available from the primary author at any time from Dr Shankar Dhakal.

## CONFLICT OF INTEREST

The authors declare no conflict of interest.

## Notes

Conflict of Interest: None

### Competing Interest Statement

The authors have declared no competing interest.

### Funding Statement

The author(s) received no specific funding for this work. NO.

### Author Declarations

Nepal Health Research Council Ref. Num. 3137 ERB Protocol Num. 263/2921P

